# First-generation clinical dual-source photon-counting CT: ultra-low dose quantitative spectral imaging

**DOI:** 10.1101/2021.11.17.21266432

**Authors:** Leening P. Liu, Nadav Shapira, Pooyan Sahbaee, Mitchell Schnall, Harold I. Litt, Peter B. Noël

## Abstract

**Objective:** Evaluation of quantification capabilities at ultra-low radiation dose levels of a first-generation dual-source Photon-Counting Computed Tomography (PCCT) compared to a dual-source dual-energy CT (DECT) scanner.

**Methods:** A multi-energy CT phantom was imaged with and without extension ring on both scanners over a range of radiation dose levels (CTDI_vol_ 0.4 - 15.0 mGy). Scans were performed in different modes of acquisition for PCCT with 120 kVp and DECT with 70/Sn150 kVp and 100/Sn150 kVp. Various tissue inserts were used to characterize the precision and repeatability of Hounsfield Units (HUs) on virtual mono-energetic images between 40 and 190 keV. Image noise was additionally investigated at ultra-low radiation dose to illustrate PCCT’s ability to remove electronic background noise.

**Results:** Our results demonstrate high precision of HU measurements for a wide range of inserts and radiation exposure levels with PCCT. We report high performance for both scanners across a wide range of radiation exposure levels with PCCT outperforming at low exposures compared to DECT. PCCT scans at lowest radiation exposures illustrate significant reduction in electronic background noise, with a mean percent reduction of 74% (p-value ∼10^−8^) compared to the 70/Sn150 kVp and 60% (p-value ∼10^−6^) compared to the 100/Sn150 kVp.

**Conclusions:** This paper reports first experiences with a clinical dual-source PCCT scanner with Quantum technology. PCCT provides reliable HUs without disruption from electronic background noise for a wide range of dose values. Diagnostic benefits are not only for quantification at ultra-low-dose but also for imaging of obese patients.

**Key Points:**

- PCCT scanners with Quantum technology provide precise and reliable quantitative Hounsfield Units at ultra-low-dose levels.
- Influence of electronic background noise can be removed at ultra-low dose acquisitions with PCCT.
- Both spectral platforms have high performance along a wide range of radiation exposure levels with PCCT outperforming at low radiation exposures.

## Introduction

After more than a decade of intensive research and development, Photon-Counting CT (PCCT) has now successfully crossed into clinical use [1]. With highly anticipated diagnostic benefits, one can foresee the replacement of current energy integrating detectors (EID) with photon-counting technology over the next decade. Early studies conducted with prototypes [2]–[11] and first clinical PCCTs [12]–[16] have illustrated significant improvements in contrast to noise ratio (CNR), spatial resolution, structural visualization, quantitative imaging, and reductions in radiation dose. These improvements became available with the drastically different detector design that enables detection of individual x-ray photons and measurement of their energies [17]–[19]. Compared to conventional CT technology, PCCT allows for improvements in clinical day-to-day routine which go beyond the availability of spectral results from every scan. One of those unique features is the potential to acquire patient scans at low radiation dose levels while offering superior quantification capabilities.

In CT, Hounsfield units (HU) represent x-ray attenuation of various tissues and different endogenous and exogenous materials. When utilizing conventional CT scanners, HUs are frequently influenced by multiple factors, including acquisition parameters, vendor-specific beam shaping (polychromatic spectrum filtration), and patient habitus. Consequently, voxel values may not represent actual tissue densities, can be ambiguous, and lack ground-truth. This limitation can theoretically be solved with use of dual-energy CT. However, the separation between high-and low-energy photons (spectral separation) in dual-energy CT can be suboptimal depending on patient habitus and employed dual-energy CT technology [20], [21]. Additionally, when imaging at ultra-low radiation dose levels, EIDs, utilized in conventional and dual-energy CTs, are challenged by electronic background noise. The electronic noise background follows a Gaussian distribution where the mean and variance reflect the dark current and readout noise of the electronics [22], [23]. At the same time, the signal statistics for a polychromatic x-ray photon spectrum follows a compound Poisson distribution. Thus, HU measurements of voxel values remain uncertain in low-dose scenarios when utilizing dual-energy technology. On the contrary, PCCT theoretically allows removal of electronic background noise. Generally, electronic background noise influences the signal detection at the lower end of the energy spectrum, and it can therefore be removed by setting low-energy PCCT signal threshold at around 25 keV [24].

The scope of this study is to characterize quantification capabilities at ultra-low radiation dose levels of a first-generation dual-source PCCT scanner (NAEOTOM Alpha, Siemens Healthineers, Forchheim, Germany) equipped with two photon-counting detectors. To illustrate improvements over latest EID technology, all experiments were performed with a PCCT and a dual-source dual-energy CT scanner (SOMATOM Force, Siemens Healthineers). We present measurements performed with a multi-energy CT phantom to characterize the precision and repeatability of HUs for various tissue types over a range of radiation dose levels. Further, we investigate the noise behavior at ultra-low radiation dose to illustrate PCCT’s ability to remove electronic background noise. Our results demonstrate the opportunity for quantitative imaging at ultra-low dose levels with PCCT, which may be key for improved diagnostics for many clinical applications, such as detection and characterization of lesions in oncology.

## Methods

### Multi-Energy CT Phantom

A multiple-energy phantom for CT performance evaluation (MECT, Sun Nuclear, Melbourne, FL, United States) (Figure 1) was imaged on two different CT systems. To evaluate the effect of patient habitus, experiments were performed with the inner phantom (20 cm diameter / small) and with the full oval shaped phantom (30 × 40 cm / large). The phantom was equipped with interchangeable tissue-simulating inserts, which, for our experiments, included: adipose, blood 70, blood 100, blood + iodine 2 mg/ml, blood + iodine 4 mg/ml, brain, calcium 50 mg/ml, iodine 2 mg/ml, iodine 5 mg/ml, and iodine 10 mg/ml. See Figure 1 for details on the corresponding position of the various inserts.

**Figure 1.**
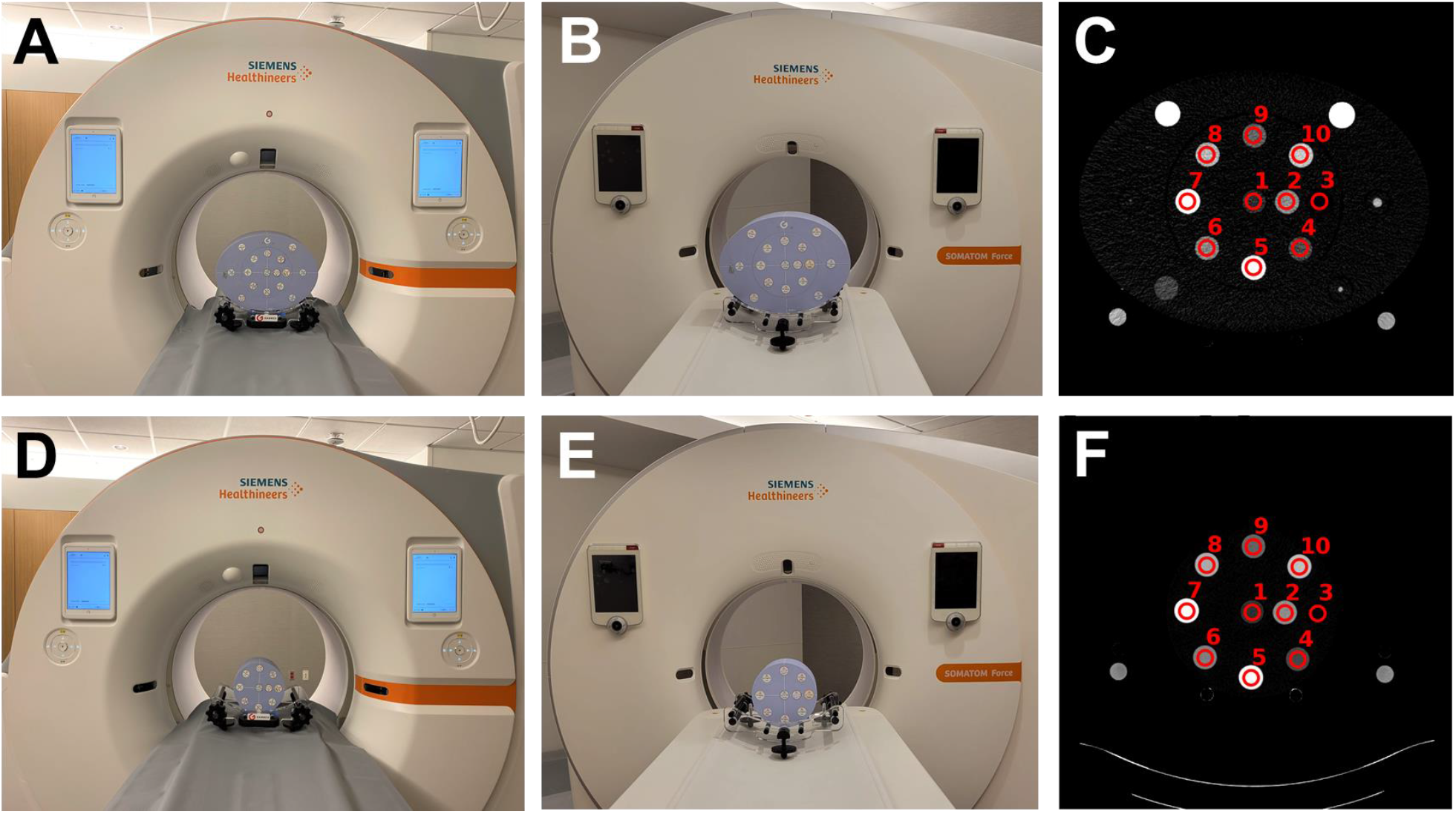
Experimental setup. (A/D) Photography of NAEOTOM Alpha and (B/E) SOMATOM Force with multi-energy CT phantom. (C/F) Reconstructed VMI 70 keV slice with numbered tissue-simulating inserts: 1. brain, 2. blood 100, 3. adipose, 4. iodine 2 mg/ml, 5. calcium 50 mg/ml, 6. blood + iodine 2 mg/ml, 7. iodine 10 mg/ml, 8. iodine 5 mg/ml, 9. blood 70, 10. blood +iodine 4 mg/ml.

### Image Acquisition & Reconstruction

Scans were performed on a first-generation dual-source PCCT with Quantum Technology (NAEOTOM Alpha, Siemens Healthineers) in QuantumPlus mode and on a dual-source dual-energy CT (SOMATOM Force, Siemens Healthineers). Dual-energy scans were performed in two different modes of acquisition: 70/Sn150 kVp and 100/Sn150 kVp. The phantom, for both sizes (small / large), was placed in the iso-center of each individual scanner (Figure 1). Data acquisition and reconstruction of the phantom was performed utilizing a standard clinical protocol (Table 1). Images were obtained, without using any exposure modulation, at multiple dose levels: CTDI_vol_ 0.4/0.6, 0.8, 1.2, 1.6, 2.0, 4.0, 6.0, 10.0, 15.0 mGy. For some dual-energy scans, the rotation time was adjusted to match PCCT dose levels. Additionally, 0.6 mGy was the minimal available dose for the 100/Sn150kVp acquisitions. Radiation dose (CTDI_vol_ values) utilized in this study convert to an effective dose range between 0.27 mSv to 10.26 mSv (k = 0.015 mSv mGy^-1^ cm^-1^) for an abdomen with a scan length of 45 cm. Each scan was repeated three times to account for the influence of statistical effects on the reconstructed results. Generated data included virtual mono-energetic images (VMI) at multiple energy levels exploiting the complete available energy range: 40, 50, 60, 70, 100, 150, 190 keV.

**Table 1.** Acquisition and reconstruction parameters

| Scanner model | NAEOTOM Alpha | SOMATOM Force |  |
| --- | --- | --- | --- |
| Tube voltage | 120 kVp (single source mode) | 70/Sn150 kVp | 100/Sn150 kVp |
| Rotation time | 0.25 | 0.25/0.5 | 0.25/0.5 |
| Spiral pitch factor | 1 | 1 | 1 |
| Collimation | 144 × 0.4 mm | 96 x 0.6 mm | 96 x 0.6 mm |
| Slice thickness | 3 mm | 3 mm | 3 mm |
| Iterative Reconstruction | QIR 3 | ADMIRE 3 | ADMIRE 3 |
| Reconstruction filter | Qr40 | Qr40 | Qr40 |
| Reconstructed field of view | 450 mm | 450 mm | 450 mm |
| Matrix size | 512 x 512 | 512 x 512 | 512 x 512 |
| Pixel spacing (in x and y) | 0.88 mm | 0.88 mm | 0.88 mm |

### Image Analysis

To evaluate quantitative stability for each VMI energy level, ten consecutive central slices were selected for analysis. Regions of interest (ROI) were placed on each individual insert on a 15 mGy 70 keV VMI for each phantom size and scanner combination. These ROIs were then copied to VMIs of other keVs and of other dose levels from the corresponding phantom size and scanner combination to measure their means and standard deviations. The mean across the three repeated scans (total of 30 slices) was subsequently calculated for each insert at each dose and VMI energy level to account for statistical effects. Analysis of quantitative repeatability with respect to the utilized radiation dose for the acquisition was performed by calculating mean HU differences relative to the HU quantifications obtained at a radiation dose level of 6.0 mGy. The relationship between relative differences in mean HU quantification and radiation dose were represented using a scatter plot for each insert and VMI energy level, where error bars represent a single standard deviation (in each direction).

Further analysis of the effect of dose on spectral quantifications was performed by evaluating root mean square errors (RMSE) across all radiation dose levels and coefficients of variation. For each insert and each keV value, RMSE was calculated relative to the mean HU value across all dose levels, while the coefficient of variation was calculated as the ratio of standard deviation to the mean CT number, i.e., attenuation quantifications relative to air (*CT#* = *HU* + 1000). Average coefficients of variation across the different material inserts, represented as percent, were plotted against VMI energy levels to demonstrate differences in variation between VMI energy levels as well as phantom size and scanner combinations.

To evaluate noise in all configurations, additional reconstructions of VMI at 70 keV were acquired without iterative reconstruction (QIR and ADMIRE off for PCCT and DECT acquisitions, respectively). Noise was computed by averaging standard deviations of ROIs from three repeated scans (30 slices) for each individual insert and radiation dose combination. Values were represented in a scatter plot, with error bars corresponding to standard deviations between slices. Characterization of the noise was achieved by utilizing the inverse square relationship between radiation dose and image noise. This relationship corresponds with Poisson noise; deviation from this relationship at lower doses indicates the presence of non-negligible system noise, such as electronic noise. Noise corresponding to scans between 1.2 and 15 mGy were linearly fit against the inverse square of their corresponding CTDI_vol_ values. RMSE values relative to predicted (linear fit) noise values for 0.4/0.6 and 0.8 mGy scans were calculated to emphasize differences from the expected theoretical relationship and visualized for each material insert in a scatter plot. Noise corresponding to scans between 1.2 and 15 mGy were linearly fit against the inverse square of CTDI_vol_. RMSE relative to predicted noise values for 0.4/0.6 and 0.8 mGy scans were calculated to emphasize differences from the theoretical relationship. RMSE for each rod were visualized in a scatter plot. A one sample t-test was utilized to examine the percent reduction in non-Poisson noise of PCCT relative to 70/Sn150 kVp pair and PCCT relative to 100/Sn150 kVp pair. A p value less than 0.05 denoted significance.

## Results

Results from the dose-dependent quantitative HU precision at 70 keV relative to the mean at CTDI_vol_ of 6.0 mGy are summarized in Figure 2. Each panel illustrates the results for the individual inserts with magnified regions for the low dose regime (CTDI_vol_ 0.4 to 5 mGy). Overall, we obtained a good agreement between the different spectral CT technologies (PCCT vs. DECT) with consistent results when comparing individual inserts. At lower dose values, considerable deviations of relative HUs can be detected for the larger patient size when imaged with DECT. Independent of patient habitus, PCCT and DECT performed similarly at higher radiation exposures, as differences due to phantom sizes became smaller with high values of CTDI_vol_.

**Figure 2.**
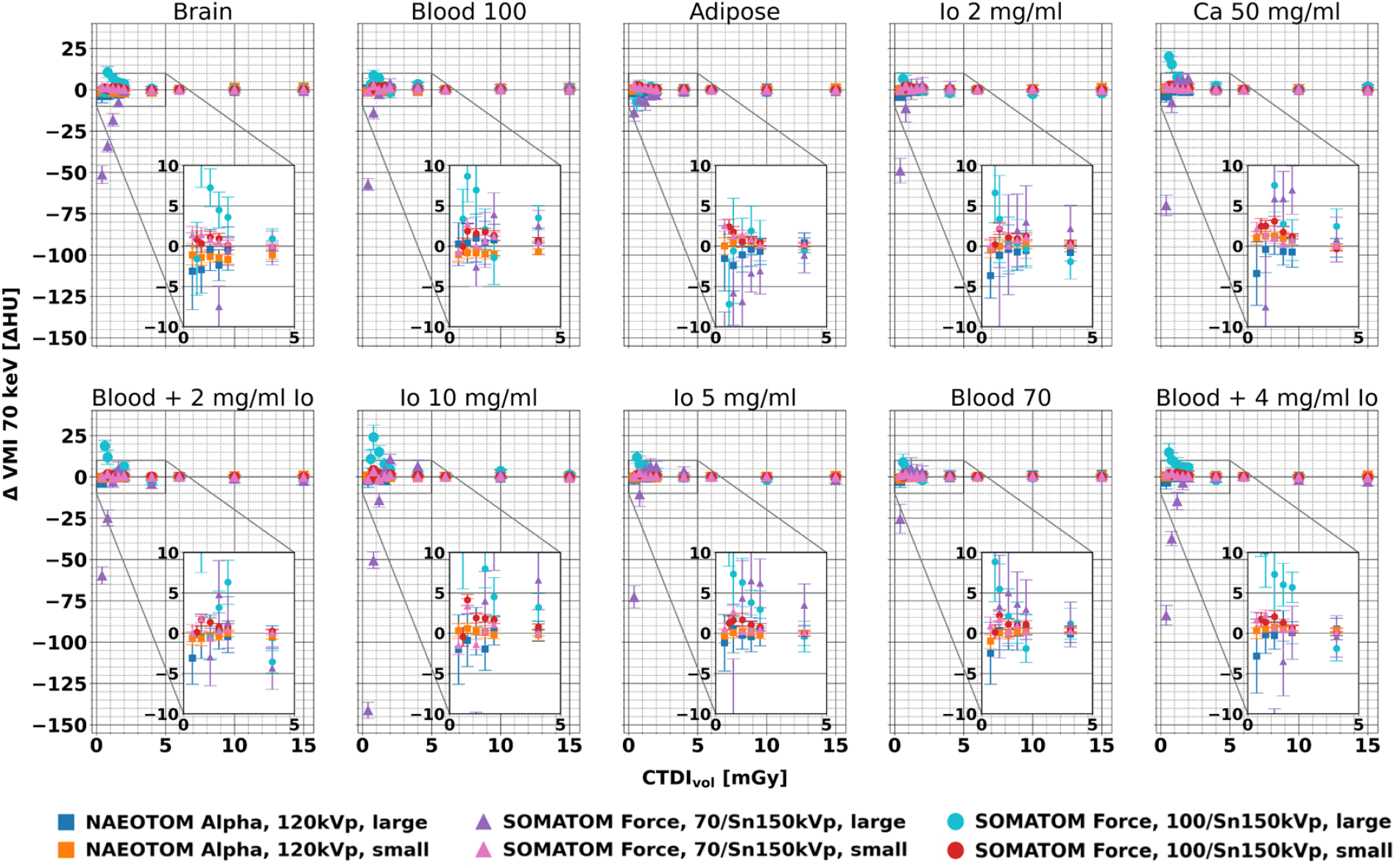
Comparison of relative HU at VMI 70 keV versus CT dose index (CTDI_vol_) for individual insert and phantom sizes between PCCT and dual-energy CT. Note that the HU differences are relative to the 6.0 mGy scan. Enlarged low-dose sections visualize regions with largest variations between scanners.

Figure 3A presents the RMSE calculated relative to the mean across all radiation dose levels for VMI 70 keV data. For this evaluation, it can be concluded that comparable results are observed across scanner platforms for each individual insert. For the larger patient size, PCCT provides reduced RMSE (0.92 ± 0.28 HU) across inserts compared to DECT with either 70/Sn150 kVp (20.32 ± 11.32 HU) or 100/Sn150 kVp (4.58 ± 1.87 HU). Coefficients of variation for each VMI energy level and scanner platform were calculated and are summarized for each insert in Table 2 (small patient size) and 3 (large patient size). Compared to earlier results where we focused on 70 keV data, in Figure 3B one can observe the average coefficient of variation for individual VMI energy levels. The previously observed trend that PCCT outpaces DECT for larger patient sizes can also be observed along the whole spectrum of VMIs.

**Table 2.** Coefficients of variation for individual inserts in large phantom.

| VMI<br>[keV] | Scanner | Tube<br>voltage<br>[kVp] | Coefficient of variation [%] |  |  |  |  |  |  |  |  |  |
| --- | --- | --- | --- | --- | --- | --- | --- | --- | --- | --- | --- | --- |
|  |  |  | Brain | Blood<br>100 | Adipose | Io 2<br>mg/ml | Ca 50<br>mg/ml | Blood + Io<br>2 mg/ml | Io 10<br>mg/ml | Io 5<br>mg/ml | Blood<br>70 | Blood + Io<br>4 mg/ml |
| 40 | NAEOTOM<br>Alpha | 120 | 0.75 | 0.63 | 0.34 | 0.78 | 0.96 | 0.95 | 0.50 | 0.92 | 0.89 | 0.86 |
|  | SOMATOM<br>Force | 70/Sn150 | 9.88 | 9.59 | 3.24 | 7.06 | 9.56 | 8.73 | 13.22 | 8.69 | 4.79 | 10.62 |
|  | SOMATOM<br>Force | 100/Sn150 | 2.08 | 0.88 | 1.76 | 0.57 | 1.07 | 1.92 | 1.21 | 0.90 | 0.88 | 0.82 |
| 50 | NAEOTOM<br>Alpha | 120 | 0.39 | 0.31 | 0.20 | 0.40 | 0.48 | 0.48 | 0.29 | 0.50 | 0.41 | 0.44 |
|  | SOMATOM<br>Force | 70/Sn150 | 5.69 | 5.54 | 1.77 | 4.39 | 5.88 | 5.39 | 9.24 | 5.73 | 2.80 | 6.79 |
|  | SOMATOM<br>Force | 100/Sn150 | 1.12 | 0.48 | 0.96 | 0.41 | 0.80 | 1.29 | 0.92 | 0.65 | 0.46 | 0.65 |
| 60 | NAEOTOM<br>Alpha | 120 | 0.17 | 0.12 | 0.13 | 0.16 | 0.17 | 0.16 | 0.12 | 0.20 | 0.13 | 0.15 |
|  | SOMATOM<br>Force | 70/Sn150 | 3.21 | 3.14 | 0.93 | 2.63 | 3.47 | 3.18 | 6.05 | 3.58 | 1.57 | 4.12 |
|  | SOMATOM<br>Force | 100/Sn150 | 0.58 | 0.30 | 0.51 | 0.31 | 0.64 | 0.88 | 0.71 | 0.48 | 0.29 | 0.53 |
| 70 | NAEOTOM<br>Alpha | 120 | 0.11 | 0.03 | 0.11 | 0.10 | 0.10 | 0.09 | 0.07 | 0.04 | 0.09 | 0.09 |
|  | SOMATOM<br>Force | 70/Sn150 | 1.71 | 1.68 | 0.44 | 1.49 | 1.90 | 1.76 | 3.68 | 2.08 | 0.82 | 2.32 |
|  | SOMATOM<br>Force | 100/Sn150 | 0.36 | 0.28 | 0.26 | 0.26 | 0.57 | 0.63 | 0.57 | 0.38 | 0.29 | 0.46 |
| 100 | NAEOTOM<br>Alpha | 120 | 0.26 | 0.18 | 0.13 | 0.34 | 0.39 | 0.39 | 0.27 | 0.38 | 0.34 | 0.40 |
|  | SOMATOM<br>Force | 70/Sn150 | 0.32 | 0.33 | 0.23 | 0.13 | 0.37 | 0.34 | 0.27 | 0.25 | 0.27 | 0.38 |
|  | SOMATOM<br>Force | 100/Sn150 | 0.54 | 0.40 | 0.21 | 0.24 | 0.51 | 0.33 | 0.48 | 0.30 | 0.45 | 0.38 |
| 150 | NAEOTOM<br>Alpha | 120 | 0.35 | 0.26 | 0.16 | 0.46 | 0.55 | 0.54 | 0.39 | 0.57 | 0.46 | 0.56 |
|  | SOMATOM<br>Force | 70/Sn150 | 1.24 | 1.20 | 0.52 | 0.90 | 1.44 | 1.29 | 2.41 | 1.42 | 0.74 | 1.71 |
|  | SOMATOM<br>Force | 100/Sn150 | 0.74 | 0.48 | 0.35 | 0.25 | 0.51 | 0.30 | 0.53 | 0.30 | 0.54 | 0.35 |
| 190 | NAEOTOM<br>Alpha | 120 | 0.37 | 0.28 | 0.16 | 0.49 | 0.59 | 0.58 | 0.42 | 0.61 | 0.49 | 0.60 |
|  | SOMATOM<br>Force | 70/Sn150 | 1.47 | 1.42 | 0.60 | 1.10 | 1.71 | 1.54 | 2.99 | 1.73 | 0.86 | 2.05 |
|  | SOMATOM<br>Force | 100/Sn150 | 0.79 | 0.50 | 0.39 | 0.25 | 0.51 | 0.32 | 0.55 | 0.31 | 0.56 | 0.35 |

**Table 3.** Coefficients of variation for individual inserts in small phantom.

| VMI<br>[keV] | Scanner | Tube<br>voltage<br>[kVp] | Coefficient of variation [%] |  |  |  |  |  |  |  |  |  |
| --- | --- | --- | --- | --- | --- | --- | --- | --- | --- | --- | --- | --- |
|  |  |  | Brain | Blood<br>100 | Adipose | Io 2<br>mg/ml | Ca 50<br>mg/ml | Blood + Io<br>2 mg/ml | Io 10<br>mg/ml | Io 5<br>mg/ml | Blood<br>70 | Blood + Io<br>4 mg/ml |
| 40 | NAEOTOM<br>Alpha | 120 | 0.31 | 0.28 | 0.60 | 0.37 | 0.51 | 0.37 | 0.12 | 0.31 | 0.38 | 0.37 |
|  | SOMATOM<br>Force | 70/Sn150 | 0.11 | 0.14 | 0.15 | 0.16 | 0.17 | 0.13 | 0.28 | 0.16 | 0.12 | 0.14 |
|  | SOMATOM<br>Force | 100/Sn150 | 0.14 | 0.29 | 0.21 | 0.16 | 0.17 | 0.13 | 0.21 | 0.11 | 0.14 | 0.16 |
| 50 | NAEOTOM<br>Alpha | 120 | 0.17 | 0.16 | 0.27 | 0.18 | 0.28 | 0.18 | 0.07 | 0.17 | 0.18 | 0.20 |
|  | SOMATOM<br>Force | 70/Sn150 | 0.05 | 0.08 | 0.10 | 0.11 | 0.08 | 0.08 | 0.20 | 0.11 | 0.07 | 0.09 |
|  | SOMATOM<br>Force | 100/Sn150 | 0.06 | 0.15 | 0.12 | 0.10 | 0.12 | 0.07 | 0.16 | 0.06 | 0.09 | 0.09 |
| 60 | NAEOTOM<br>Alpha | 120 | 0.11 | 0.09 | 0.09 | 0.05 | 0.13 | 0.05 | 0.04 | 0.06 | 0.07 | 0.08 |
|  | SOMATOM<br>Force | 70/Sn150 | 0.04 | 0.07 | 0.09 | 0.08 | 0.05 | 0.06 | 0.14 | 0.08 | 0.05 | 0.07 |
|  | SOMATOM<br>Force | 100/Sn150 | 0.03 | 0.09 | 0.09 | 0.07 | 0.10 | 0.05 | 0.12 | 0.05 | 0.07 | 0.06 |
| 70 | NAEOTOM<br>Alpha | 120 | 0.11 | 0.07 | 0.03 | 0.04 | 0.04 | 0.04 | 0.02 | 0.02 | 0.04 | 0.02 |
|  | SOMATOM<br>Force | 70/Sn150 | 0.05 | 0.08 | 0.09 | 0.07 | 0.07 | 0.06 | 0.11 | 0.07 | 0.05 | 0.07 |
|  | SOMATOM<br>Force | 100/Sn150 | 0.05 | 0.07 | 0.09 | 0.07 | 0.11 | 0.05 | 0.11 | 0.07 | 0.06 | 0.07 |
| 100 | NAEOTOM<br>Alpha | 120 | 0.16 | 0.10 | 0.17 | 0.15 | 0.12 | 0.16 | 0.05 | 0.13 | 0.13 | 0.13 |
|  | SOMATOM<br>Force | 70/Sn150 | 0.08 | 0.11 | 0.10 | 0.07 | 0.12 | 0.08 | 0.09 | 0.08 | 0.07 | 0.10 |
|  | SOMATOM<br>Force | 100/Sn150 | 0.09 | 0.09 | 0.12 | 0.09 | 0.13 | 0.09 | 0.10 | 0.10 | 0.08 | 0.12 |
| 150 | NAEOTOM<br>Alpha | 120 | 0.19 | 0.13 | 0.24 | 0.21 | 0.19 | 0.22 | 0.08 | 0.19 | 0.18 | 0.20 |
|  | SOMATOM<br>Force | 70/Sn150 | 0.10 | 0.12 | 0.11 | 0.08 | 0.15 | 0.09 | 0.12 | 0.10 | 0.08 | 0.13 |
|  | SOMATOM<br>Force | 100/Sn150 | 0.11 | 0.12 | 0.13 | 0.10 | 0.15 | 0.10 | 0.11 | 0.12 | 0.08 | 0.15 |
| 190 | NAEOTOM<br>Alpha | 120 | 0.20 | 0.14 | 0.26 | 0.23 | 0.21 | 0.23 | 0.09 | 0.20 | 0.19 | 0.21 |
|  | SOMATOM<br>Force | 70/Sn150 | 0.11 | 0.12 | 0.11 | 0.08 | 0.15 | 0.10 | 0.13 | 0.10 | 0.09 | 0.13 |
|  | SOMATOM<br>Force | 100/Sn150 | 0.12 | 0.13 | 0.13 | 0.11 | 0.15 | 0.11 | 0.12 | 0.13 | 0.09 | 0.15 |

**Figure 3.**
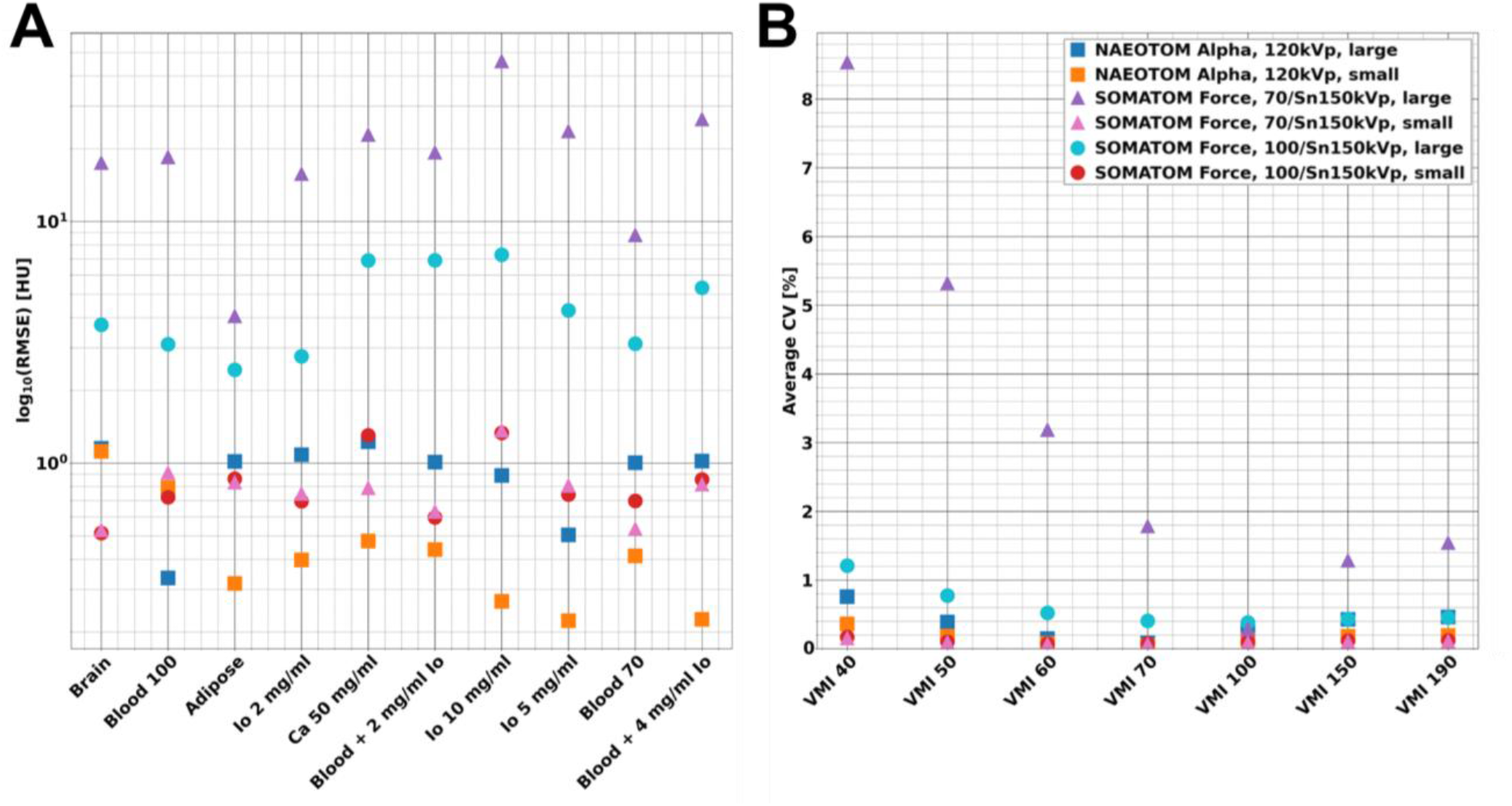
Comprehensive overview of dose dependent spectral HU quantification for PCCT and dual-energy CT. RMSE over all radiation exposure levels and VMIs versus individual inserts (A). Average coefficient of variation including all dose levels and inserts versus different VMIs (40 to 190 keV) (B). PCCT performance is marginally influenced by patient size when comparing to dual-energy CT.

Image noise was evaluated as the standard deviation within an ROI for each of the material inserts. Figure 4 presents noise dependence on radiation dose level, phantom size, spectral technology, and acquisition type at VMI 70 keV. At high and medium radiation dose levels, comparable noise levels were observed for all spectral technologies and acquisition types per each phantom size. For the large phantom size, a transition occurs at lower radiation dose levels where the PCCT technology outperforms the dual-energy technology (both kVp pairs). To further analyze this transition, we analyzed the 70 keV noise dependence as a function of inverse square root of the radiation dose, as shown in Figure 5. It is expected that for Poisson (quantum) distributed statistical events the noise has a linear dependance. Indeed, linear fits to the medium and high radiation dose levels (higher or equal to 1 mGy) demonstrate a high level of correlation, with R^2^-values of 0.9575, 0.9976, and 0.9907, for the PCCT, 70/Sn150 kVp pair, and 100/Sn150 kVp pair, respectively. Deviations from the expected linear correlation indicate the non-negligible influence of electronic noise. Figure 5B presents RMSE values of noise assessments for the two lowest radiation dose levels (0.4 and 0.8 mGy for PCCT and the 70/Sn150 kVp pair, 0.6 and 0.8 mGy for the 100/Sn150 kVp pair) across all material inserts. We observed a significant reduction in non-Poisson, i.e., electronic, noise for the PCCT scans, with mean percent noise reduction of 74% (p-value 3.33 × 10^−8^) compared to the 70/Sn150 kVp pair acquisitions and 60% (p-value 8.91 × 10^−6^) compared to the 100/Sn150 kVp pair acquisitions.

**Figure 4.**
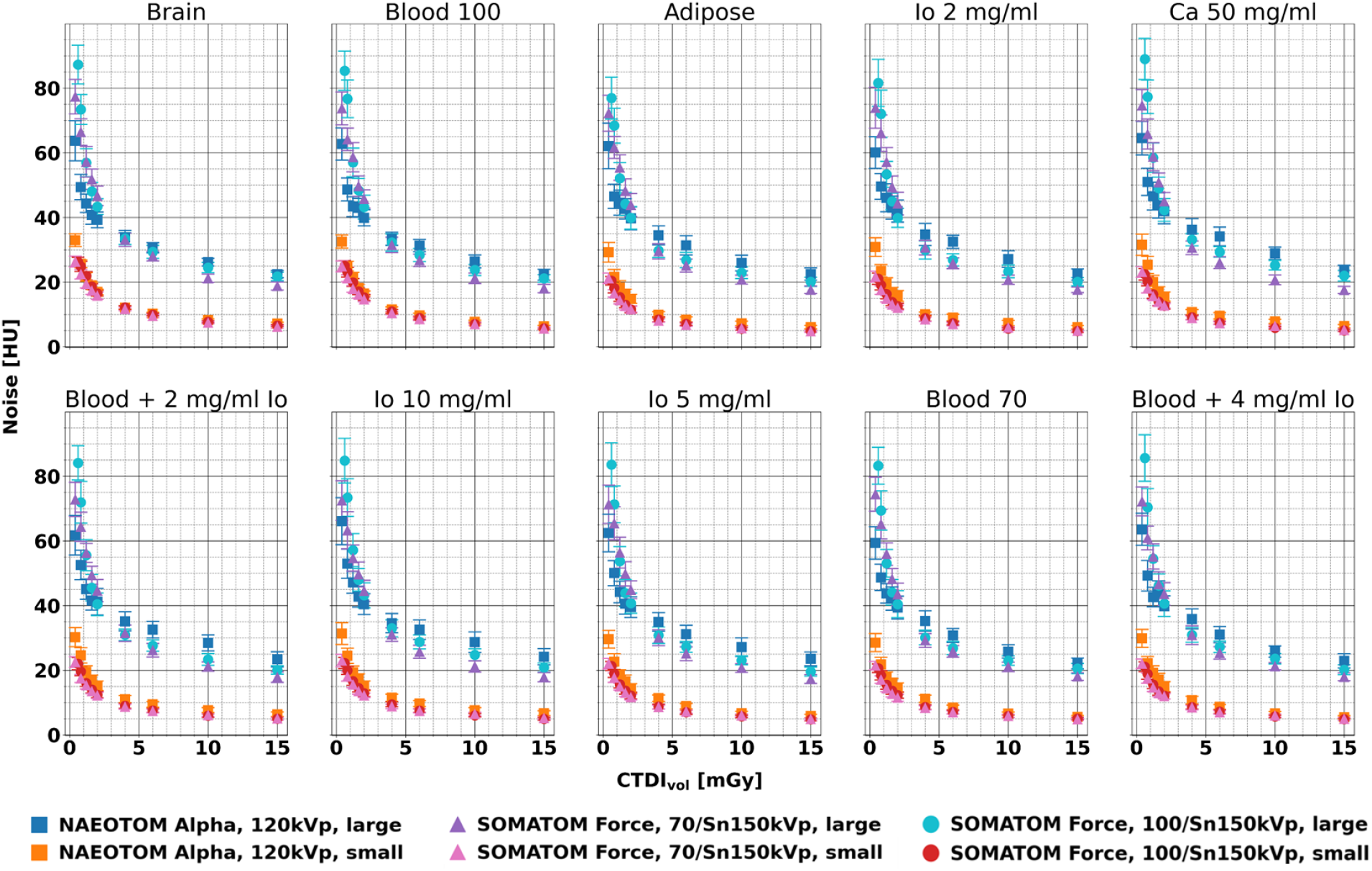
Comparison of noise levels at VMI 70 keV versus CT dose index (CTDI_vol_) for individual insert and phantom sizes between PCCT and dual-energy CT. While comparable noise levels were observed for all scanners at high and medium radiation dose levels, at lower radiation dose levels PCCT outperforms the dual-energy technology for the larger patient size.

**Figure 5.**
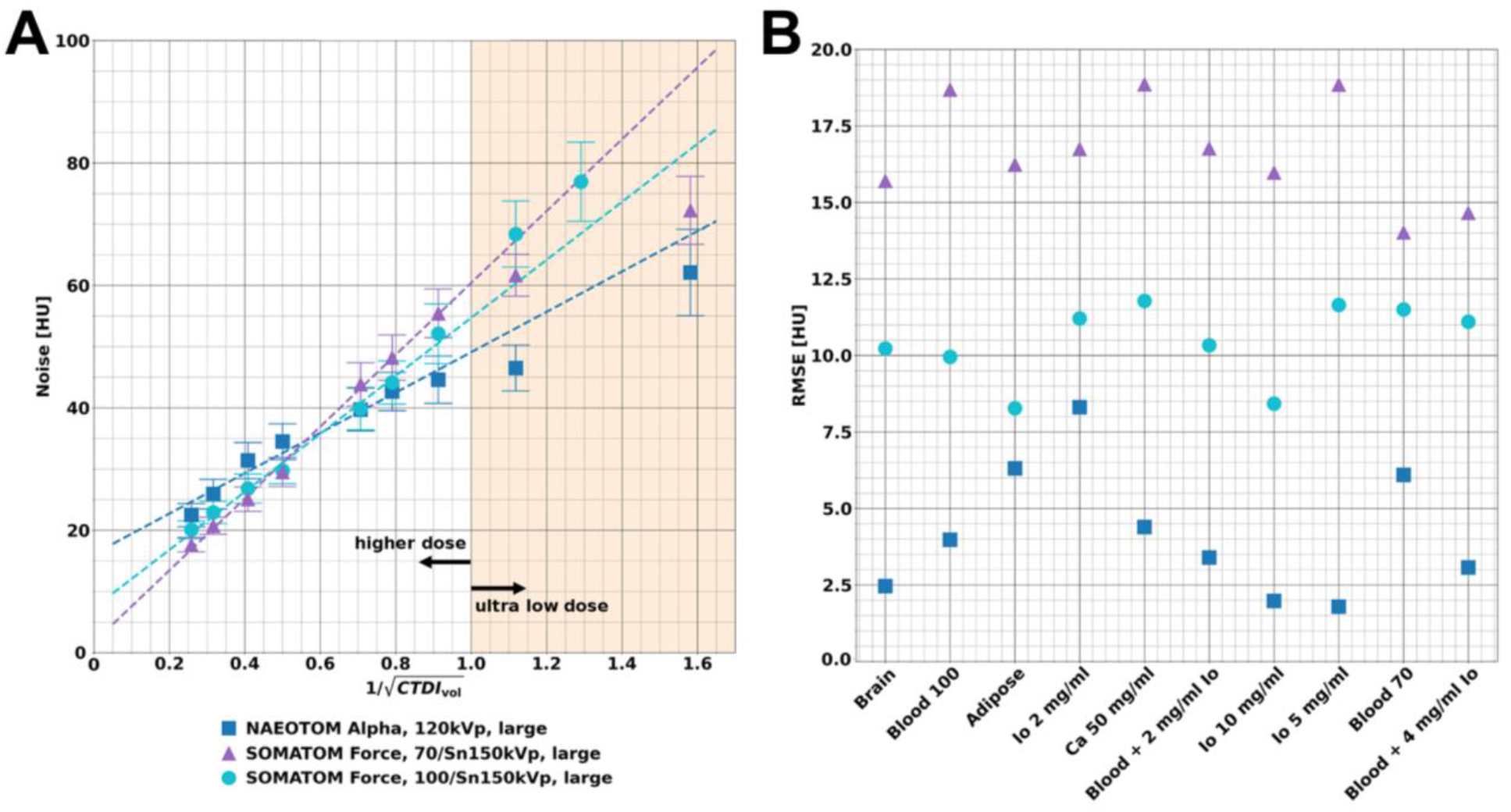
Analysis of noise dependance on radiation dose using linear fits and deviations from expected behavior from Poisson (quantum) distributed events. (A) Example of the noise dependance on radiation dose of a single material insert (adipose) and linear fits using all CTDI_vol_ **≥** 1 mGy data points (white area). Noise from low CTDI_vol_ (**<**1 mGy) data points (orange area) were not included in the linear fits. (B) Deviations from linear fits of the two lowest dose levels for different material inserts under different spectral acquisition modes. PCCT exhibits the lowest deviations, implying a small contribution of non-Poisson, e.g., electronic, noise sources.

## Discussion

This paper reports the first experience with a clinical dual-source CT scanner with Quantum Technology on spectral HU quantification and noise behavior at ultra-low-dose levels. The presented results illustrate unique features which become available with clinical PCCT: **(i)** high precision of HU measurements for a wide range of inserts and radiation exposure levels, and **(ii)** reduced influence of electronic background noise at ultra-low dose acquisitions. We report high performance for both scanners along a wide range of radiation exposure levels with PCCT outperforming the latest generation dual-energy CT at low exposures. Our data demonstrate that PCCT offers diagnostic benefits not only for quantification at ultra-low dose but also for imaging of morbidly obese patients. PCCT’s potential to reduce artifacts, such as rings, when imaging large patients has been previously demonstrated with a prototype scanner [25]. With the arrival of PCCT in the clinical arena, more accurate characterization of tissues and enhanced perfusion imaging at reduced radiation dose levels become routinely available.

Over the last decade, several investigators have reported on low-dose and/or quantitative imaging potentials of PCCT. *Gutjahr et al*. reported, with a PCCT prototype, on noise and CNR behavior for clinically relevant dose levels and found similar noise behavior with improved CNR compared to EID acquisitions [8]. *Leng et al*. demonstrated that high accuracy for iodine quantification (RMSE of 0.5 mg/ml) and accurate CT numbers in VMIs (percentage error of 8.9%) can be achieved in a range of CTDI_vol_ between 9.1 to 42.9 mGy [10]. Several groups have demonstrated the capabilities of low-dose PCCT for thoracic applications with the consensus that high spatial resolution imaging is feasible within a clinically accepted dose range [9], [26]–[28]. Concerning quantitative PCCT imaging, *Symons et al*. evaluated the effect of beam hardening on head and neck CT angiography. Their results demonstrated that patient-habitus related effects, such as beam hardening, can be eliminated with PCCT, which allowed significantly improved HU accuracy [4]. With respect to K-edge imaging, *Si-Mohamed et al*. illustrated that PCCT can non-invasively determine the biodistribution of gold nanoparticles with high correlation to optical emission spectrometry [29]. These and several other contributions used prototype systems and played a significant role in the clinical translation by illustrating significant improvements of PCCT compared to CT equipped with EIDs. Our current work demonstrates potential diagnostic advantages with a first-generation clinical PCCT by focusing on the combination of ultra-low dose and quantitative spectral imaging.

The present study has limitations. Due to the vast increase in parameter space with PCCT, we only present an initial overview of ultra-low-dose capabilities. Future studies will be necessary to evaluate additional parameters such as iterative reconstruction level, contrast agent concentrations, and effect of high spatial resolution modes on quantitative PCCT performance. Concerning contrast agents, we have not evaluated the quantitative performance concerning K-edge agents, such as nanoparticles [30]. The feature to detect and quantitatively measure K-edge agents is essential to distinguish between two contrast agents, which is required for dual-contrast protocols [2], [31]–[37]. Our experiments were only performed with a geometric image quality phantom which lacks textures seen in clinical CT acquisitions. Ultimately, findings will be confirmed in the clinical routine, but as an intermediate step, we plan to utilize patient-based phantoms [38]. Finally, our study only includes the comparison to one dual-energy CT scanner. Other generations or implementation of dual-energy CT or conventional EID CT may provide different performance in the utilized radiation dose range.

## Conclusion

In conclusion, the present study reports on experimental evaluations of a first generation dual-source PCCT scanner with a focus on quantitative imaging at ultra-low radiation dose levels. Compared to conventional CT and DECT, PCCT provides reliable HUs without disruption from electronic background noise for a wide range of dose values and patient sizes. This may lead to a paradigm change for the clinical day-to-day routine with enhanced capabilities and increased confidence for a wide range of diseases.

## Data Availability

All data produced in the present study are available upon reasonable request to the authors.

## Acknowledgment

We acknowledge support through the National Institutes of Health (R01EB030494).

